# Lupus RGMX: Social and Clinical Characteristics and their Contribution to Quality of Life in a Mexican Cohort with SLE

**DOI:** 10.1101/2023.02.23.23286331

**Authors:** Hernández-Ledesma Ana Laura, Martínez Domingo, Fajardo-Brigido Elizabeth, Talía V. Román-López, Nuñez-Reza Karen, Vera del Valle Sandra Valentina, Domínguez-Zúñiga Donaji, Tinajero-Nieto Lizbet, Peña-Ayala Angélica, Torres-Valdez Estefania, Frontana-Vázquez Gabriel, Gutierrez-Arcelus Maria, Rosetti Florencia, Alcauter Sarael, Miguel E. Rentería, Alejandra E. Ruiz-Contreras, Alpízar-Rodríguez Deshiré, Medina-Rivera Alejandra

## Abstract

**BACKGROUND:** Although higher prevalence, disease activity, damage accumulation and mortality of systemic lupus erythematosus (SLE) are observed among Latin American, North American admixed population, African descendants and Native Americans, the information about SLE in Latin American countries, such as Mexico, is scarce.

**OBJECTIVES:** To present Lupus RGMX, a multidisciplinary effort to generate a national digital patient registry to enrich the understanding of Mexican people with SLE.

**METHODS:** Mexican patients with SLE registered between May 2021 and January 2023 in Lupus RGMX were included. Sociodemographic, socioeconomic, and clinical characteristics, along with quality-of-life perception (QoL) were assessed using self-reported data. We compared the QoL obtained from patients with SLE with two groups of non-SLE Mexican subjects. Descriptive statistics, comparisons analyses and a multivariate nonparametric regression model were performed.

**RESULTS:** A total of 1172 of lupus patients were included; of which 93.9% were women. The mean age±SD was 36.6±10.7 years, with 37.1% of the individuals between 41 and 50 years. The 24.9% reported a calculated monthly income of 430 USD (8,612 MXN). Lower QoL scores were observed in the SLE group, especially in subjects with lower socioeconomic level. Health perception, QoL perception and socioeconomic status were the variables with greater importance to predict total WHOQoL scores.

**CONCLUSION:** The design and implementation of Lupus RGMX imply a pioneering approach to unraveling SLE in Mexicans. Further studies from Lupus RGMX will be focused on enriching the representation of the Mexican population and include other aspects that may allow us to improve our understanding of the disease in our population.

## 1. INTRODUCTION

Patient registries enable the creation of large databases with demographic and clinical data for patients with a specific disease. Their implementation can provide valuable information about the onset and progression of the disease, its clinical features, epidemiology, course of treatment, clinician decision-making, healthcare services utilization and cost of treatment; therefore, these databases are highly valuable for medical research and public health policies (McGettigan et al., 2019).

Systemic Lupus Erythematosus (SLE) is a multi-systemic autoimmune disease with a widely heterogeneous set of clinical manifestations (Goulielmos et al., 2018). Previous studies have shown the influence of ethnicity and socioeconomic status on SLE’s risk and prognosis. Higher disease activity, organ damage accumulation, as well as a higher rate of mortality has been observed in SLE patients from Latin American, North American admixed population, African descendants, and Native American patients (Barber et al., 2021; Drenkard & Lim, 2019; Lewis & Jawad, 2017; Mendoza-Pinto et al., 2022).

The application of classification criteria, such as the American College of Rheumatology (ACR) and the European League Against Rheumatism (EULAR), alongside improving the quality and scope of national health systems, has enabled the design and implementation of adequate registries of patients with SLE (Aringer et al., 2019; Lewis & Jawad, 2017). Unfortunately, existing registries and studies are mainly focused on European and European descendants’ populations, which limits the adequate representation of minorities, such as Latin-Americans, and hinders the generalization and accessibility of the obtained knowledge (Drenkard & Lim, 2019; Izmirly et al., 2021). For instance, the National Lupus Patient Registry of the United States represents one of the largest of these initiatives and includes well known studies (CDC, 2022). For populations considered as Hispanic there are two important references: the Lupus Registry of the Spanish Society of Rheumatology (RELESSER) and the Grupo Latino Americano de Estudio del Lupus (GLADEL). The GLADEL is a multicenter initiative that has focused for more than 20 years on improving the understanding of the characteristics and needs of Latin-American people with SLE, highlighting the clinical and social relevance of studying how SLE affects people through different countries in Latin-America. To date, GLADEL group has achieved the integration of a longitudinal cohort of Latin-American individuals with SLE that has allowed them to evaluate potential biomarkers, risk factors, treatments, and clinical features specific to our populations, representing a landmark in the field (Gómez-Puerta et al., 2021; B. A. Pons-Estel et al., 2004; Rúa-Figueroa et al., 2014). The GLADEL and GLADEL 2.0 cohort included 248 and 145 Mexican individuals, respectively, from 7 different centers in México City, Guadalajara, San Luis Potosí and Monterrey (Gómez-Puerta et al., 2021; B. A. Pons-Estel et al., 2004). Despite efforts have been made in order to improve our knowledge about SLE in Latin-America, there is still scarce information about it in México; to date, there are no comprehensive epidemiological reports on the prevalence and incidence and, until recently, there was no epidemiological surveillance system or registry collecting data from patients across national medical institutions to enable research and the identification of their specific characteristics and unmet needs (Gómez-Puerta et al., 2021).

Here we present an overview of the methodology followed for the establishment of Lupus RGMX, the Mexican Lupus Registry, a digital data collection platform designed to deepen into the knowledge and understanding of the characteristics of the Mexican people with SLE and do a follow up along the years. Through the implementation of Lupus RGMX and its digital availability, we have launched a pioneering approach for the understanding of SLE in México.

## 2. METHODS

### 2.1. Ethics statement

This project was reviewed and approved by the Ethics on Research Committee of the Institute of Neurobiology at Universidad Nacional Autónoma de México (UNAM). Personal and clinical information were anonymized and stored at the National Laboratory of Advanced Scientific Visualization at UNAM. At the beginning of the evaluation, participants provided informed consent, and a copy of the privacy statement was given to each volunteer, in accordance with the Federal Law on the Protection of Personal Data Held by Private Parties.

### 2.2. Patient selection criteria and recruitment

Participants were recruited on a voluntary basis; initially through social media campaigns (i.e., Facebook and Instagram) along with a website with information about the project (https://lupusrgmx.liigh.unam.mx/). Additionally, we reached out to some of the biggest and most active lupus foundations and patient support groups in Mexico (Fundación Proayuda Lupus Morelos A.C and Lupus MX) and established alliances. Furthermore, formal invitations to participate in the recruitment of patients have been extended to rheumatology specialists in Mexico. Eligible participants must have a current diagnosis for SLE by a rheumatologist, be aged over 18 years, be able and willing to provide informed consent and complete the study questionnaires. Open recruitment for this analysis was held between May 2021 and January 2023. Disease activity and comorbidities were not considered as exclusion criteria.

### 2.3. Design of Lupus RGMX project identity

For the Mexican Lupus Registry’s acronym, the word *Lupus* was selected as the central element; followed by the letters *RG*, from ReGistry and *MX*, from MeXican. The study’s logo integrates a butterfly, which resembles the malar rash, one of the most common manifestations of SLE, along with a meaning of hope; the purple color palette was selected because of its wide utilization in awareness campaigns as it symbolizes a middle point between the intensity and motivation of the red and the calmness and stability of the blue.

The website of the project (https://lupusrgmx.liigh.unam.mx/) was designed using the same color palette as in the logo.

### 2.4. Database collection platform

A digital data collection platform was implemented using the Research Electronic Data Capture (REDCap) hosted at UNAM (Harris et al., 2009, 2019). REDCap provides a secure and friendly web-based platform for both the design and application of surveys and for the subsequent generation and management of the data. The availability of online questionnaires facilitates reaching people throughout the national territory, which favors having a more homogeneous and representative sample.

### 2.5. Survey and instruments

Four questionnaires were included, covering (i) the collection of general and sociodemographic data, (ii) the evaluation of socioeconomic status, (iii) clinical manifestations and medical history and (iv) the assessment of quality of life. All questionnaires were suitable for self-application.

#### 2.5.1. Sociodemographic characteristics

The first questionnaire included the informed consent and privacy statement acceptance. Personal information such as name, gender, date and place of birth were required fields and were used as filters to avoid duplicated registries. Existence of a previous lupus diagnosis and consumption of SLE specific treatments were included in this survey in order to classify participants as either individuals with SLE or controls.

#### 2.5.2. Socio-economic status

The socioeconomic status of the registered individuals was determined using the socioeconomic level index (NSE 2022) rule of the Mexican Association of Market and Opinion Intelligence Agencies (AMAI) (*NSE*, 2022). The NSE is an algorithm used to classify Mexican homes based on the evaluation of six dimensions of household welfare: (i) human capital, (ii) practical infrastructure, (iii) connectivity and entertainment, (iv) health infrastructure, (v) planning/future and (vi) availability of basic infrastructure and space. It includes 6 items with different scores values, with a total of 300 points able to be obtained.

The NSE contemplates seven socioeconomic levels: A/B (202+ points), C+ (168-201), C (141-167), C- (116-140), D+ (95-115), D (48-94) and E (0-47). A higher score implies a better capacity to fulfill the necessities within a household (*NSE*, 2022).

#### 2.5.3. Clinical manifestations and medical history

The clinical questionnaire was only accessible for participants with a previous lupus diagnosis, and it includes items about clinical characteristics associated with SLE (on the basis of rheumatologist recommendations), treatment, pregnancy, presence of comorbidities and familiar history of lupus. Additionally, data from health service providers was included. Due to the educational and socioeconomic heterogeneity of our population and, in order to ensure an easier and more accessible participation that provides more realistic and meaningful data, we designed infographics that explained in plain and non-technical language each of the clinical manifestations included in our surveys. These infographics were reviewed and approved by both rheumatologist and presidents of lupus patient associations in México and were therefore integrated in the online platform.

#### 2.5.4. Quality of life

To assess quality of life (QoL) we used the Spanish Spain version of the World Health Organization Quality of Life Questionnaire (WHOQOL-BREF). It contains 26 items validated in a 5-point Likert scale; 24 from WHOQOL-100 questionnaire and two additional items to evaluate overall quality of life and general health perception. Higher scores are indicative of a better QoL (*The World Health Organization*, n.d.). A profile of QoL perception was generated; environmental, physical, psychological, and social domains were evaluated, as well as overall quality of life and health perception; this profile was compared with two groups of non-SLE subjects: the first group was integrated using non-SLE individuals registered on the Lupus RGMX platform, invited directly by the SLE participants and through social media. The second group was identified from the Mexican Registry of Twins (Twins MX) a registry previously established by the Neurogenomics consortium, of which Lupus RGMX is part, that shares the design and surveys with Lupus RGMX (Leon-Apodaca et al., 2019); for this analysis only one individual per family was considered.

#### 2.5.5. Data initial management and statistical analysis

To avoid possible duplicated registries, the database was filtered. Based on name, cell phone number and email address, an automated script was implemented in R 4.2.2 (R Core Team, 2022) to find duplicates, in such cases the completeness among duplicated registries were compared and chosen the completest one. It is worth mentioning that, to keep anonymity, the script ran blinded (i.e, without printing participants names). Additionally, as Lupus RGMX is a self-reported registry it is possible that we have some false lupus registries, in accordance with the Lupus RGMX’s rheumatologists’ suggestions, we established a filter for classical treatments corresponded to SLE including glucocorticoids, antimalarials, Mycophenolic acid, Azathioprine, Methotrexate, Rituximab, Cyclosporine and Cyclophosphamide. Thus, every participant consuming at least one treatment type was included in further analysis. Besides, we removed registries with atypical ages, i.e., ranging around one thousand years, surely due to typing mistakes.

For the descriptive statistics analysis bar plots and box plots were produced using ggplot2 package (Wickham, 2009), the maps with the distribution of SLE subjects and rheumatologists across Mexico were made using mxmaps (Valle-Jones D, 2022) and intersecting sets of clinical manifestations were visualized using UpSetR package (Conway et al., 2017).

Statistical analysis consisted of three parts; the first one compared the WHOQoL scores of SLE-individuals with two groups of non-SLE individuals, the second part compared the WHOQoL scores only from SLE-individuals grouped by socioeconomic level, and the third part explored the marginal effects of some variables over Total-WHOQoL score. Next, we show a detailed description for each part.

First, to find out if the quality of life is different in SLE-group (n = 1214, 1138 women, mean-age = 37.06, SD = 38.48), we compared the overall score WHOQoL, as well as the scores of environmental, physical, psychological, and social domains to two independent groups of non-SLE individuals (**Supplementary Table S1**). One comparison group came from volunteers that completed the Lupus RGMX registry by RedCap as healthy controls (n = 179, 132 women, mean-age = 30.56, SD = 11.55), from here called the RedCap volunteers group. The other comparison group came from volunteers who participated in the TwinsMX registry (n = 1271, 924 women, mean age = 29.29, SD = 49.04), from here called the Twin volunteers group. Since TwinsMX project had the same WHOQoL questionnaires in Mexican population and capture the sames sociodemographic variables in multiple-births healthy individuals in the same temporality as Lupus RGMX, we be able to compare them to SLE-group, taking only one individual per family. After dropping out of uncompleted registries, we kept 1578 cases; 942 from SLE-group (888 women), 128 from RedCap volunteers group (87 women), and 508 from Twins volunteers group (395 women).

Since we faced an intrinsic imbalance issue in all the samples (i.e., more women with SLE than men, and disproportionate size samples), we implement 200 random resamples (Templ, 2016) to compute the independent permutation test (Fay, 2022) of our variables of interest among each dyad of groups. Based on age distribution and sex proportion, we paired each resample and computed the mean difference and its significance. Each resample was size n = 100 with 90% of women. After computing 200 resamples, we reported the expected result. Our program analysis was developed and implemented in R 4.2.2 language (R Core Team, 2022), which is available on https://github.com/NeuroGenomicsMX/Lupus_RGMX_analysis.

Secondly, to find out if the total quality of life changes in SLE-individuals across different socioeconomic levels, we considered only the SLE-group, and evaluated socioeconomic-status total score and WHOQoL total score completeness. After dropping out of uncompleted registries, we kept 966 cases (906 women). Next, following AMAI rules, we transformed the numerical total score from socioeconomic status to categories. It is worth mentioning that categories C+ and A/B were empty, which implies the cohort does not have any SLE participant from the highest socioeconomic levels. Furthermore, as we mentioned before, cases with atypical ages were removed. Then, total WHOQoL scores across socioeconomic levels E, D, D+, C-, and C were compared by Kruskal-Wallis rank sum test to figure out if any differences exist, later we applied a pairwise permutation test (Mangiafico, 2023) to find where those differences belong to. Such tests were corrected by a false discovery rate method.

Finally, to explore the effect of some variables on Total-WHOQoL score (target variable), we applied a multivariate adaptive regression splines model (Milborrow, 2023) to the SLE-group data set. Predictor variables were, overall QoL perception, general health perception, socioeconomic status, calculated age, sex, years living with SLE, prednisolone, treatment, comorbidity, health-service provider, and diagnostic lag. Target, as well as predictor variables, are described in the **Supplementary Table S2**.

Prednisolona, Treatment, Comorbidity, and health provider were transformed to dummy variables by one-hot encoding technique, no other preprocessing was applied to the data set. After dropping out of uncompleted registries and filter atypical ages, we kept 712 cases, with 45 predictor variables and total-WHOQoL as target variable (**Supplementary Table S2**).

The model implemented 15 cross validation replicas (Kuhn et al., 2022) and, once the best tune converged, the relative importance of variables was computed (Greenwell, B et al., 2022), finally, marginal effects were calculated for the most important variables (Greenwell, 2022).

## 3. RESULTS

### 3.1. Sociodemographic characteristics

One thousand two hundred and fourteen registries were recovered at the Lupus RGMX digital platform from May 2021 to January 2023. For descriptive analysis, inconsistent and incomplete registries were eliminated, recovering a total of one thousand one hundred and seventy-two registries, with a mean age of 36.6 (±10.7) years. Almost ninety-four percent were women (**see Supplementary Table S3**). Nearly half of the participants were employed (41.4%), whereas regarding education level the patients that had a college degree were 37,7%, followed by those with a senior high school or technician degree (26.4%). We identified that 9 (0.8%) of the women included in the registry were pregnant, 263 (22.4%) of the participants reported a previous diagnosis of lupus nephritis and 211 (18%) having a family member with SLE. Almost fifty-seven percent of the patients were being attended at IMSS or at ISSSTE, the biggest public health institutions in México, whereas 29.9% were in private institutions. Besides glucocorticoids (91.2%), antimalarials were the most frequent treatment among the patients (27.5%).

The mean age of the cohort was 36.6 ± 10.7 years, ranging from 18 to 79 years, with the majority (37.11%) between 41 and 50 years (**Figure 1A**); duration of the disease was from 1 month to 62 years, with a mean of 8.6 ± 7.9 years (**Figure 1B**), with a mean age at diagnosis of 28.1± 10.8 years, ranging from 2 to 74 years, with the majority (39.2%) being diagnosed at 31-40 years (**Figure 1C**).

**Figure 1.**
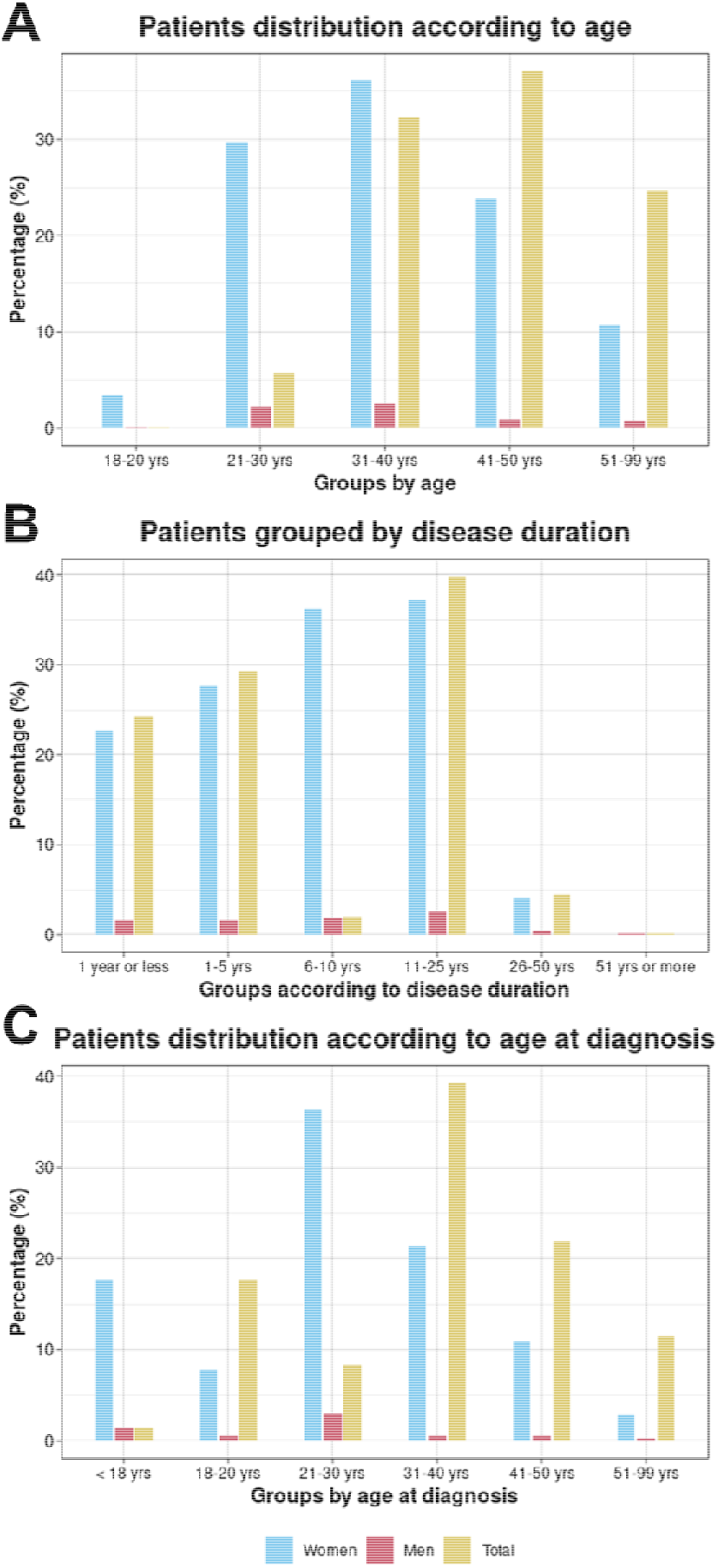
Distribution according to (A) age, (B) disease duration and (C) age at diagnosis in the participants with Systemic Lupus Erythematosus (n=1172).

Registries from the 32 states of Mexico were collected through Lupus RGMX (**Figure 2, see supplementary table 2**). More than 50% of the registries come from 4 states: México City (n=235), State of México region (n=131), Querétaro (n=107) and Morelos (n=60); whereas the states with less representation were Aguascalientes (n=3), Baja California Sur (n=2) and Colima (n=2).

**Figure 2.**
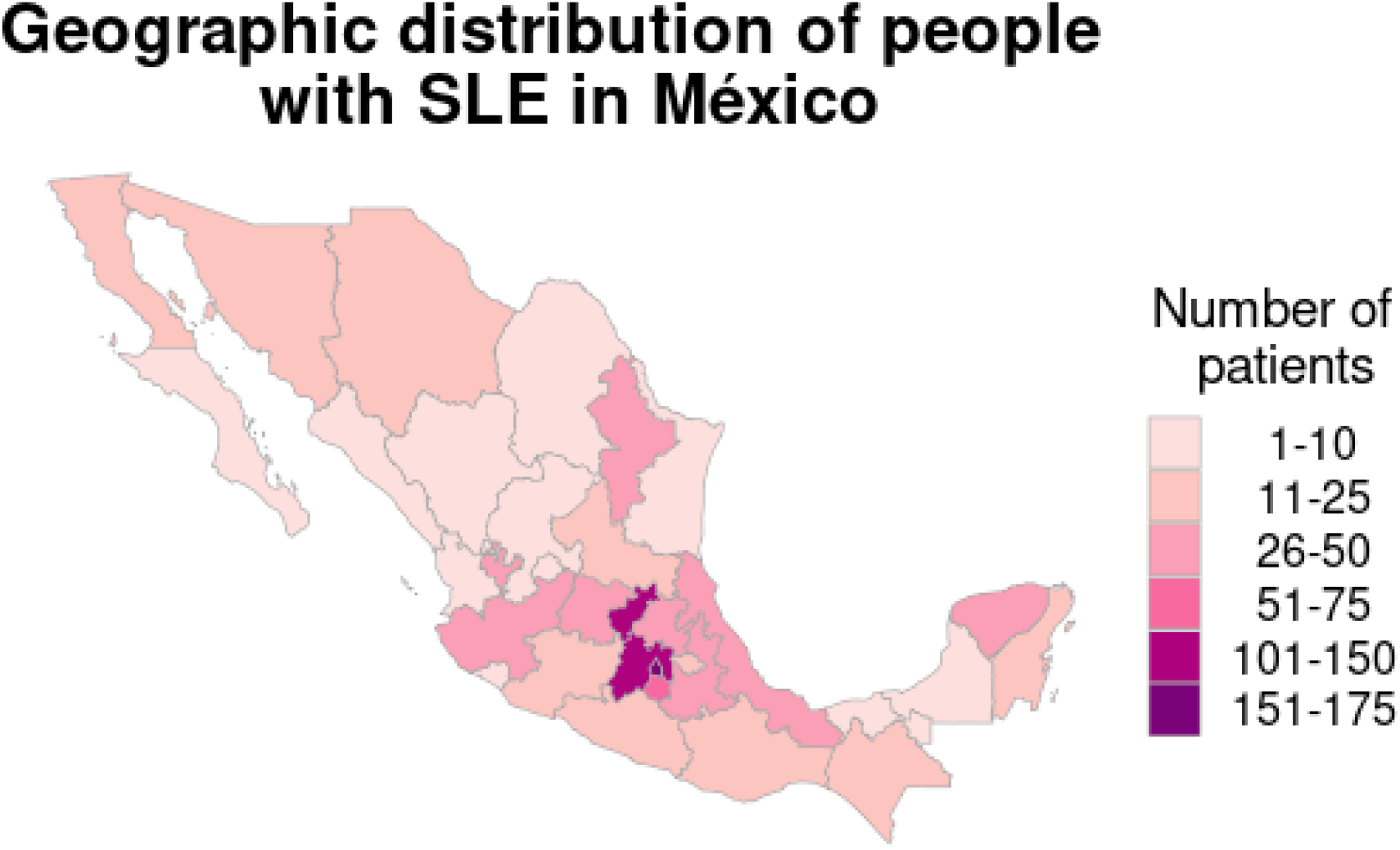
Geographic distribution of individuals with Systemic Lupus Erythematosus (n=1172).

### 3.2. Socio-economic status

Participants were distributed among the five lower levels established by NSE, with the majority between levels E and D (**Figure 3**). The highest level observed was the D level, this implies a monthly home mean income of 8,156 MXN (approx. 430 USD) or less for all the individuals, according to the National Household Expenditure Income Survey, ENIGH (INEGI, 2020). Still, it is important to highlight that the majority (75%) were among the E (24.9%) and the D (22.8%) levels, which implies a monthly income between 4,950 and 11,285 MXN (around 261 and 595 USD, respectively).

**Figure 3.**
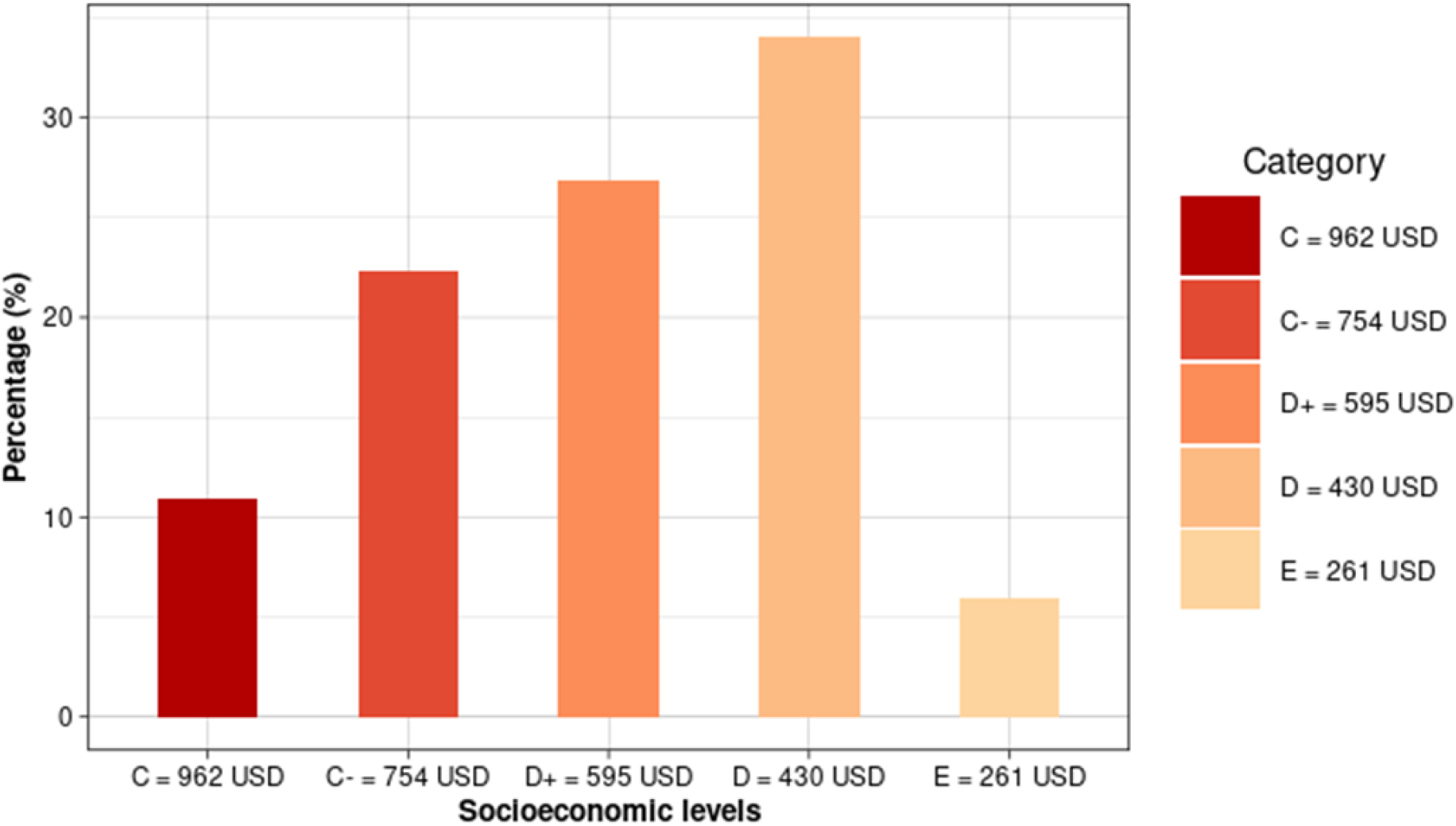
Socioeconomic level distribution of individuals with Systemic Lupus Erythematosus (n=1172).

### 3.3. Clinical manifestations and medical history

Hypertension (12%) and thyroid diseases (11%) were the most frequent comorbidities among SLE patients, whereas forty-two percent did not have any comorbidity (**Figure 4A**). Arthritis (68%) and alopecia (59%) were reported as the most frequent manifestations, followed up by lupus headache (44%) and mucosal ulceration (35%); whereas increased urinary casts (7%), seizure (5%), and pyuria (1%) were the least frequent manifestations (**Figure 4B**). Afterwards, we analyzed the co-occurrence of mild and severe manifestations, according to SLEDAI criteria, across patients (**Figure 4C**); individual total frequency of each manifestation, gray and purple bars on the bottom left, showed arthritis and lupus headache as the most frequent manifestations as observed on figure 4B. Frequency of co-occurrence of manifestations, represented as intersects in the matrix on the bottom right, showed that co-occurrences involving arthritis are the most frequent.

**Figure 4.**
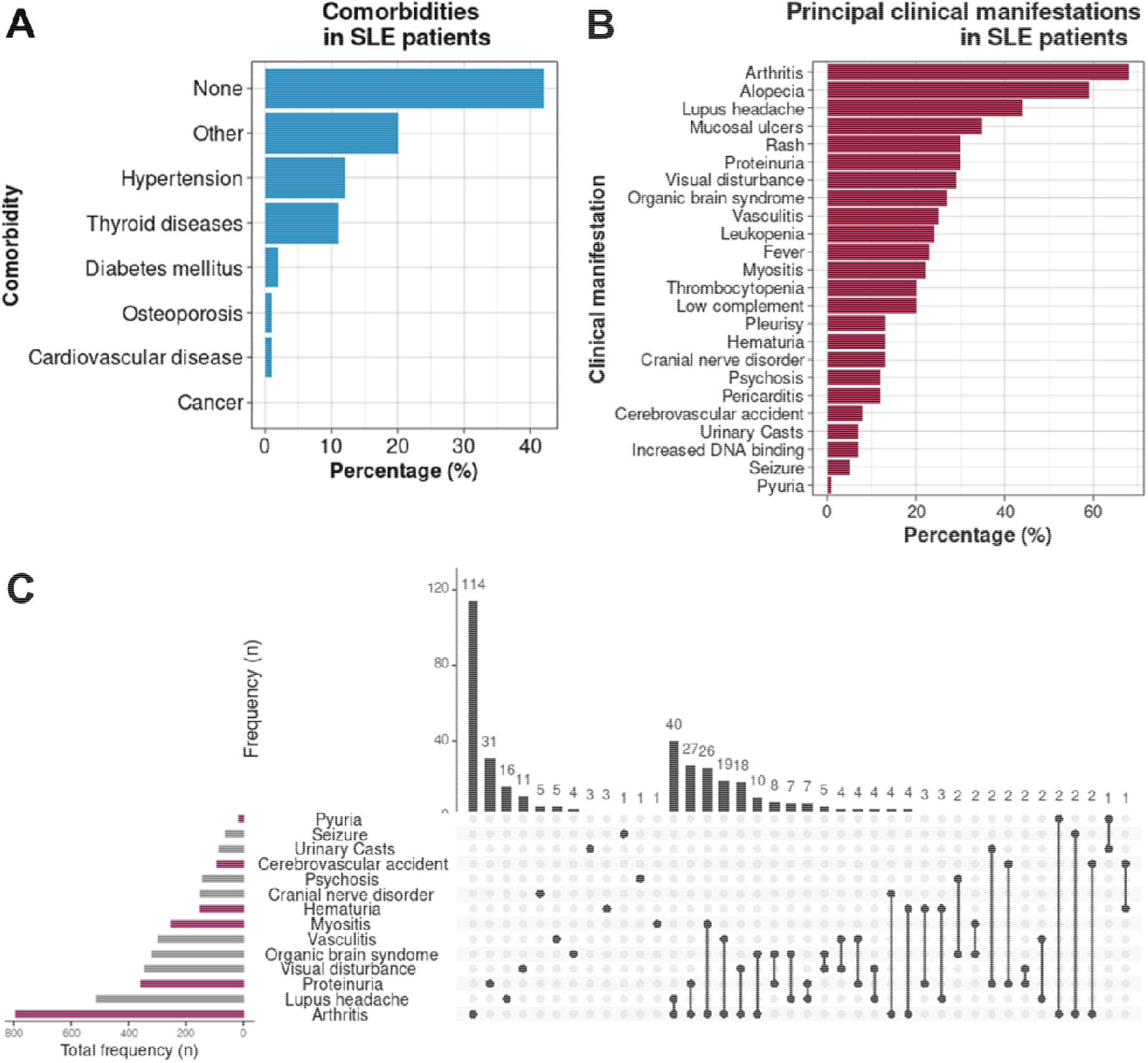
Self-reported clinical manifestations by the individuals registered in Lupus RGMX (n=1172). Frequency of comorbidities (A), clinical manifestations (B) and co-occurrence of manifestations (C).

### 3.4. Quality of life analysis

#### 3.4.1. SLE v.s. non-SLE individuals

Regarding total WHOQoL scores, permutation tests between SLE-patients and RedCap volunteers, as well as, between SLE-patients and Twins volunteers showed significant differences. Specifically, SLE-group scored 7.67 points lower than RedCap volunteers (p-value = 0.000); and SLE-group also scored 10.49 points lower than Twins volunteers (p-value = 0.000). In other words, SLE individuals have the lowest scores of total WHOQoL as can be seen in **Figure 5**.

**Figure 5.**
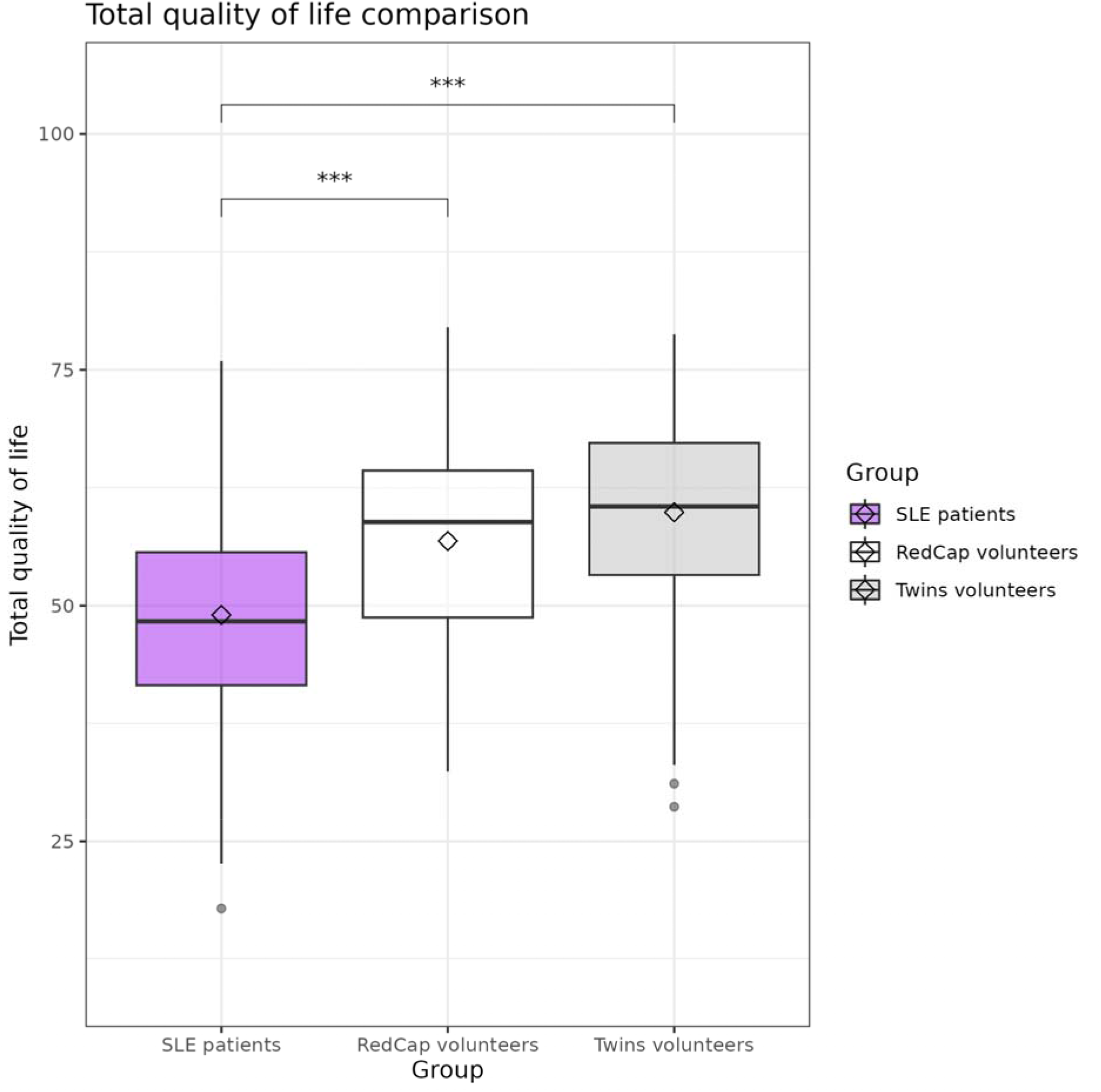
Total WHOQoL score differences among SLE-patients (n = 942) and two groups of non-SLE (RedCap volunteers n = 128, TwinsMX volunteers n = 395). Regarding overall WHOQoL, RedCap volunteers scored 7.67 (SE = 1.26) higher than SLE patients (p-value = 0.000), whereas Twins volunteers scored 10.49 points (SE = 1.21) above SLE patients (p-value = 0.000). The SLE-group showed the lowest QoL.

When we consider scores in four domains of WHOQoL, permutation test results indicate significant differences among all groups in all domains (see **Supplementary Table S5**). Figure 6 shows significant scores differences among groups ordered by specific domain, namely, panel (A) environmental domain, panel (B) physical-health domain, panel (C) psychological domain, and panel (D) social-relations domain. For all QoL domains, we found significant differences between SLE patients and RedCap volunteers, as well as between SLE patients and Twins volunteers. In all cases, the SLE group scored the lowest.

**Figure 6.**
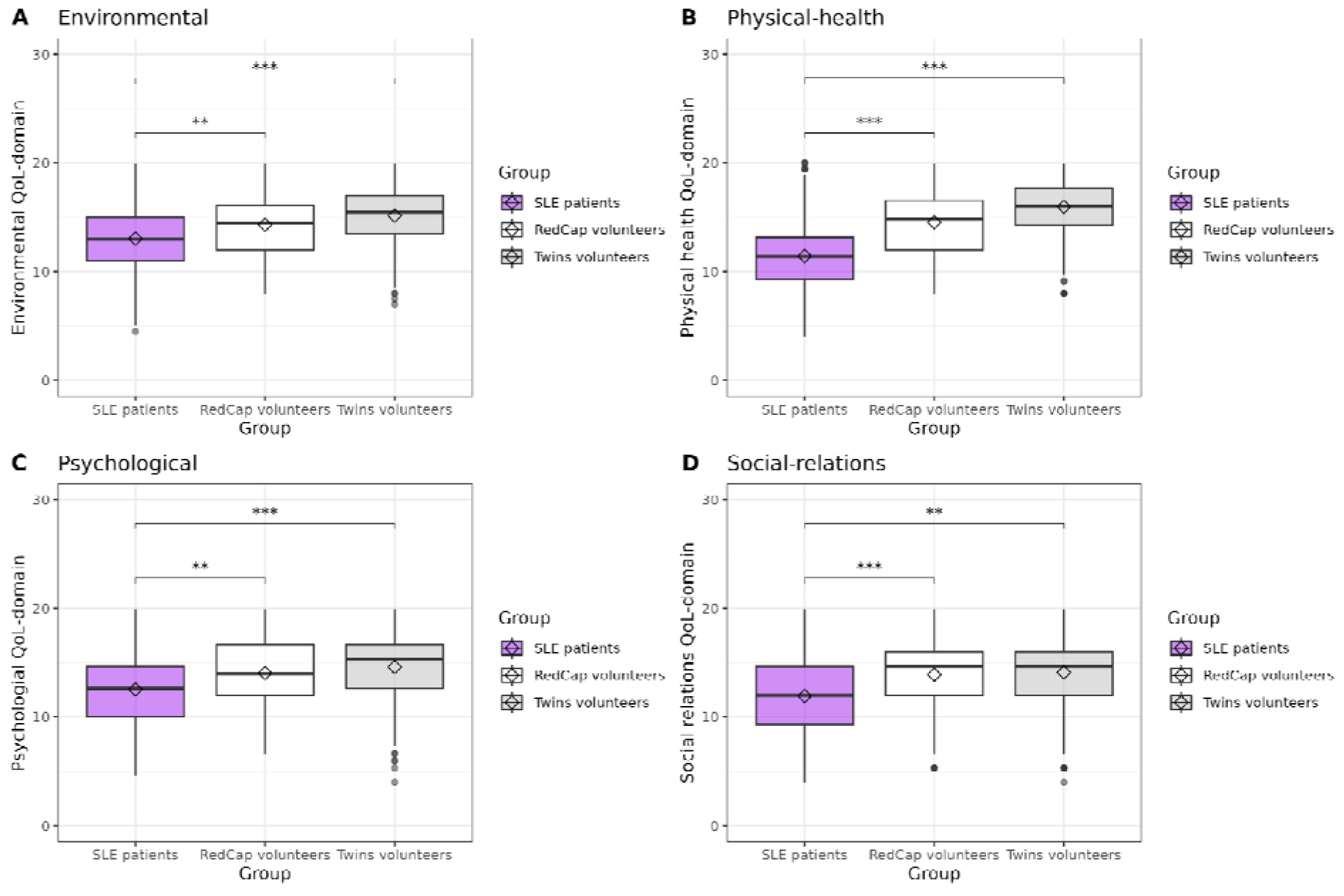
WHOQoL scores differences among SLE-patients (n = 942) and two groups of non-SLE individuals, i.e., RedCap volunteers (n = 128), and TwinsMX volunteers (n = 395). Each panel corresponds to one domain: A) Environmental, B) Physical-health, C) Psychological, and D) Social domain.

For the environmental domain, SLE-patients scored on average 1.12 and 2.10 lower than RedCap and Twins-group, respectively. Regarding physical-health domain, SLE-group scored 2.89 and 4.30 points below RedCap and Twins respectively. About psychological domain, SLE individuals rated 1.61 and 2.18 points lower than RedCap and Twins severally. And, for social-relations domain, SLE-group punctuated 2.13 and 2.01 below RedCap and Twins-group, respectively. Concisely, the QoL score of SLE-patients is always the lowest in all four domains.

#### 3.4.2. Socioeconomic level and Total WHOQoL in SLE individuals

To search out if any difference exists in the total quality of life of SLE patients across socioeconomic levels, we applied the non-parametric test of Kruskal-Wallis. Since the result of the Kruskal-Wallis rank sum test indicates that at least one significant difference among economic levels exists (p-value = 0.000), we ran the pairwise permutation test to find out where that difference, or differences, exists. Such results are shown visually in Figure 7 (numeric values can be seen in **Supplementary Table S6**): socioeconomic levels are presented from lowest E (261 USD) to the highest exhibited in our cohort C (962 USD); it is important to highlight that except for D and D+, contiguous levels didn’t show significant differences. On the other hand, all the noncontinuous levels showed significant differences. Average punctuation increases monotonously as the economic level grows.

**Figure 7.**
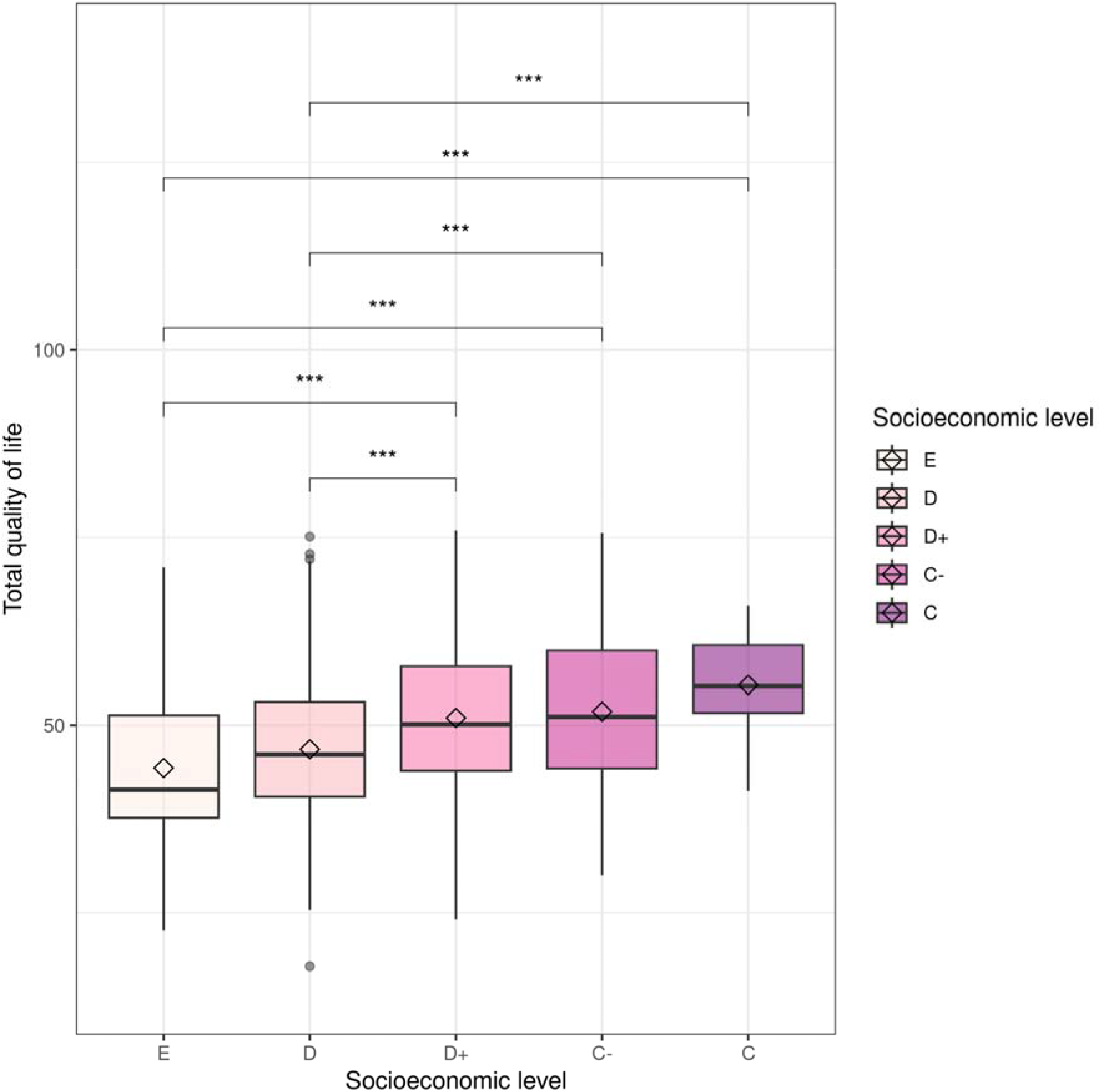
Comparison of total QoL score of individuals with SLE (n = 966) grouped by socioeconomic level. All p-values were adjusted by false discovery rate.

After correcting p-values by false discovery rate technique, we observed significant differences in total QoL along no consecutive levels, ranging from 2 up to almost 6 points of QoL. Furthermore, average punctuation increases monotonously as the economic level grows. We can also see that dispersion into the highest economic level is narrow. And it is important to mention that there are no any participants belonging to levels C+ and A/B. This is an intrinsic bias in the present study.

#### 3.4.3. Partial effects on Total WHOQoL in SLE individual

To evaluate effects over Total WHOQol of some variables of interest, we use the variables described in **Supplementary Table S2** as predictors to build a multivariate adaptive regression splines model.

After tuning the model (search grid) by cross validation (15 replicas), the optimal model included only first-degree interactions and kept five predictor variables, namely, health perception, quality of life perception, socioeconomic status, corticosteroid, and diagnostic lag. The assessing performance parameters for the optimal model were, RMSE = 7.939 points of total WHOQoL, R^2^ = 0.390, and MAE = 6.325.

Then, based on RSS value, we estimated the relative importance for the previously mentioned retained variables. As shown in **Figure 8,** importance order goes like this, health perception, QoL perception, socioeconomic status, corticosteroid and, finally, diagnostic lag. Even though some variables look mildly influential, all of them exert a certain effect over the total WHOQoL. However, it is important to emphasize that perception-type variables, such as health perception and QoL perception, are the most important variables for predicting the WHOQoL score in SLE-patients.

**Figure 8.**
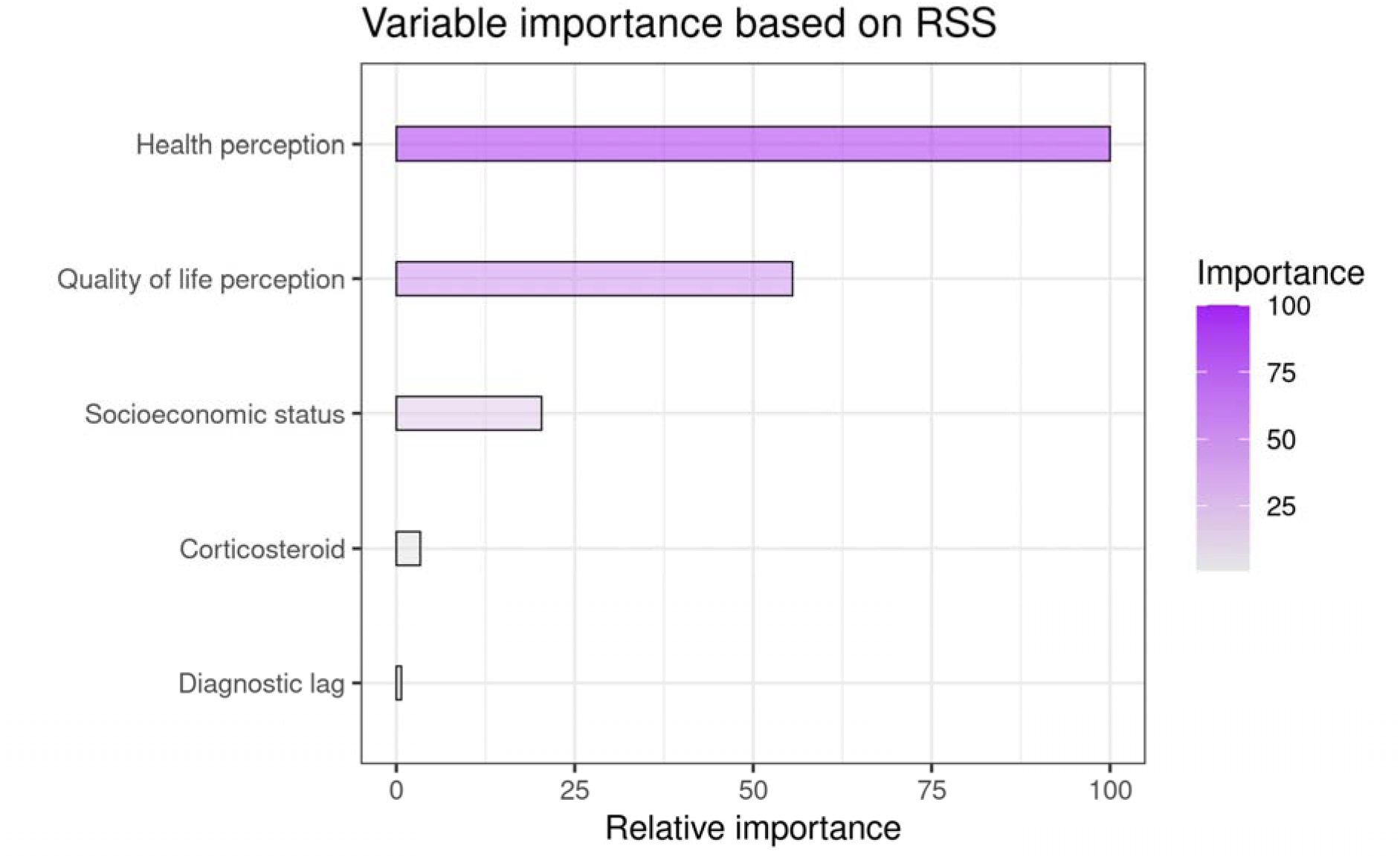
Importance of predictor variables.

Once we identified the best predictor variables, and their respective importance, we evaluated their partial (marginal) effect exerted over the total WHOQoL score. As we can see in Figure 9; when health perception rates over 4 points, there is a light marginal increase on the total QoL score (**Figure 9A**); when quality of life perception scores around 4 points, and exceed it, a sharply marginal-increase in the total QoL is observer (**Figure 9B**); if socioeconomic score rates from 0 to around 112 points, total QoL increase at constant rate but, when exceeding 112 points, there is no growth for total QoL (**Figure 9C**). If SLE patients take corticosteroids, their total QoL falls by approximately 1.7 points (**Figure 9D**). Finally, when the diagnostic lagged less than approximately 1 year, the QoL was raised by almost 2 points (**Figure 9E**); ideally early diagnosis must be done during the first year since symptoms onsets, later diagnostics have no effects on QoL. It is also worth mentioning that socioeconomic status affects QoL up to 112 points (socioeconomic level D+), beyond such level there is no effect exerted over QoL (**Figure 9C**).

**Figure 9.**
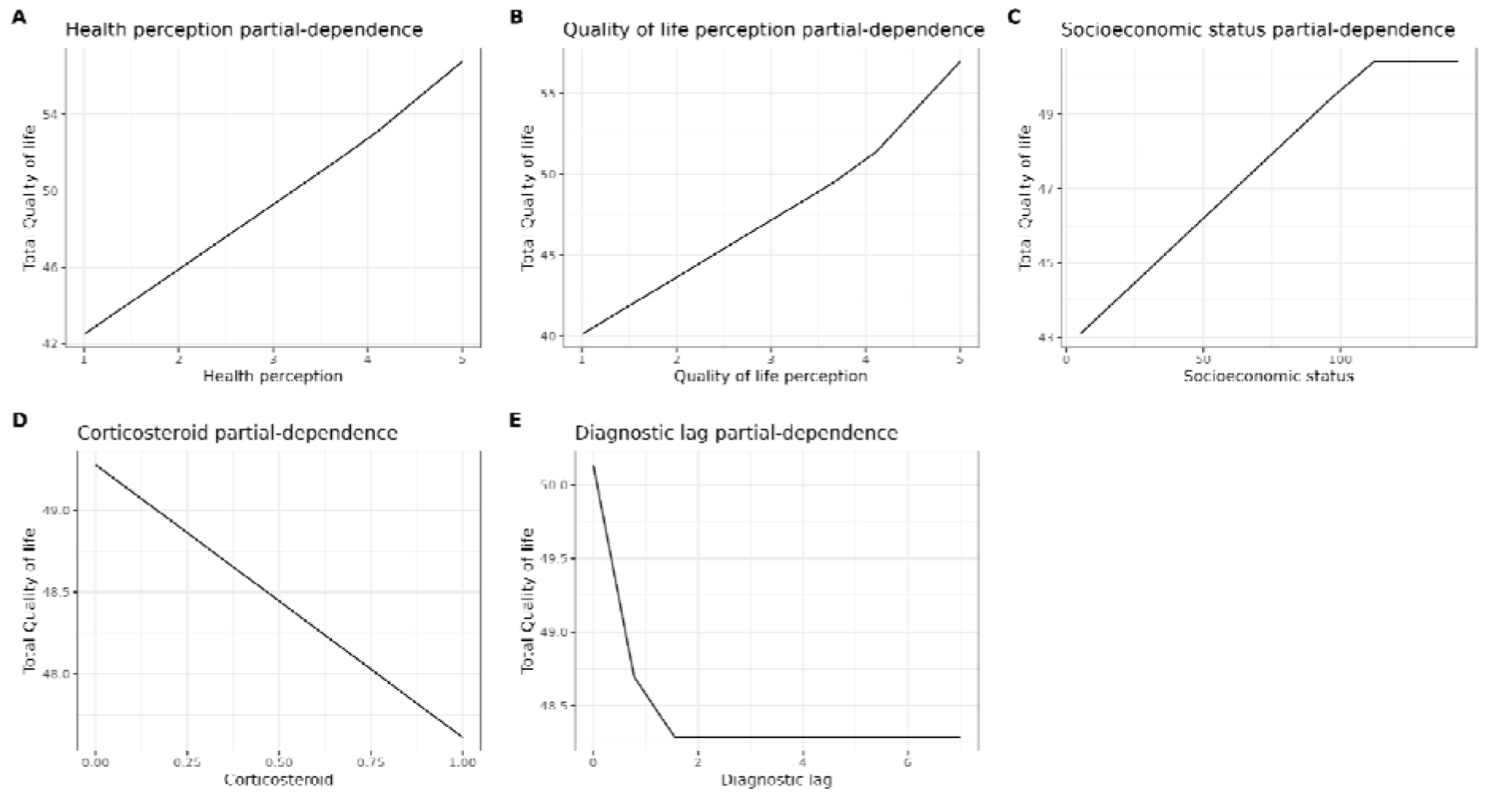
Partial dependence plots. Marginal effect that important variables exert over total WHOQoL. Whereas health perception (panel A) and taking-corticosteroids (D) showed almost linear effects; QoL perception (panel B), socioeconomic status (panel C), and diagnostic lag (panel E) showed no linear effects.

To observe the combined effects of two variables over total WHOQol, we show interaction plots of selected pairs of variables. When combined, health-perception and QoL-perception marginal effects, we can appreciate an acute augmentation in the Total QoL score, ranging from near 35 to more than 60 points (**Figure 10A**), this combination of variables produces the strongest effect of all possible combinations. In **Figure 10B** and **Figure 10D** we can see that early SLE diagnosis can diminish the negative effect associated with taking corticosteroids and lower socioeconomic levels, respectively, which highlight the importance of an early SLE diagnostic. Now, in **Figure 10C** we appreciate how socioeconomic status almost eliminates the effect of taking corticosteroids, meaning that a higher socioeconomic level could compensate for the negative effects of taking corticosteroids. Finally, in **Figure 10D** it is important to note how an early SLE-diagnostic boost QoL scores along the whole socioeconomic spectrum.

**Figure 10.**
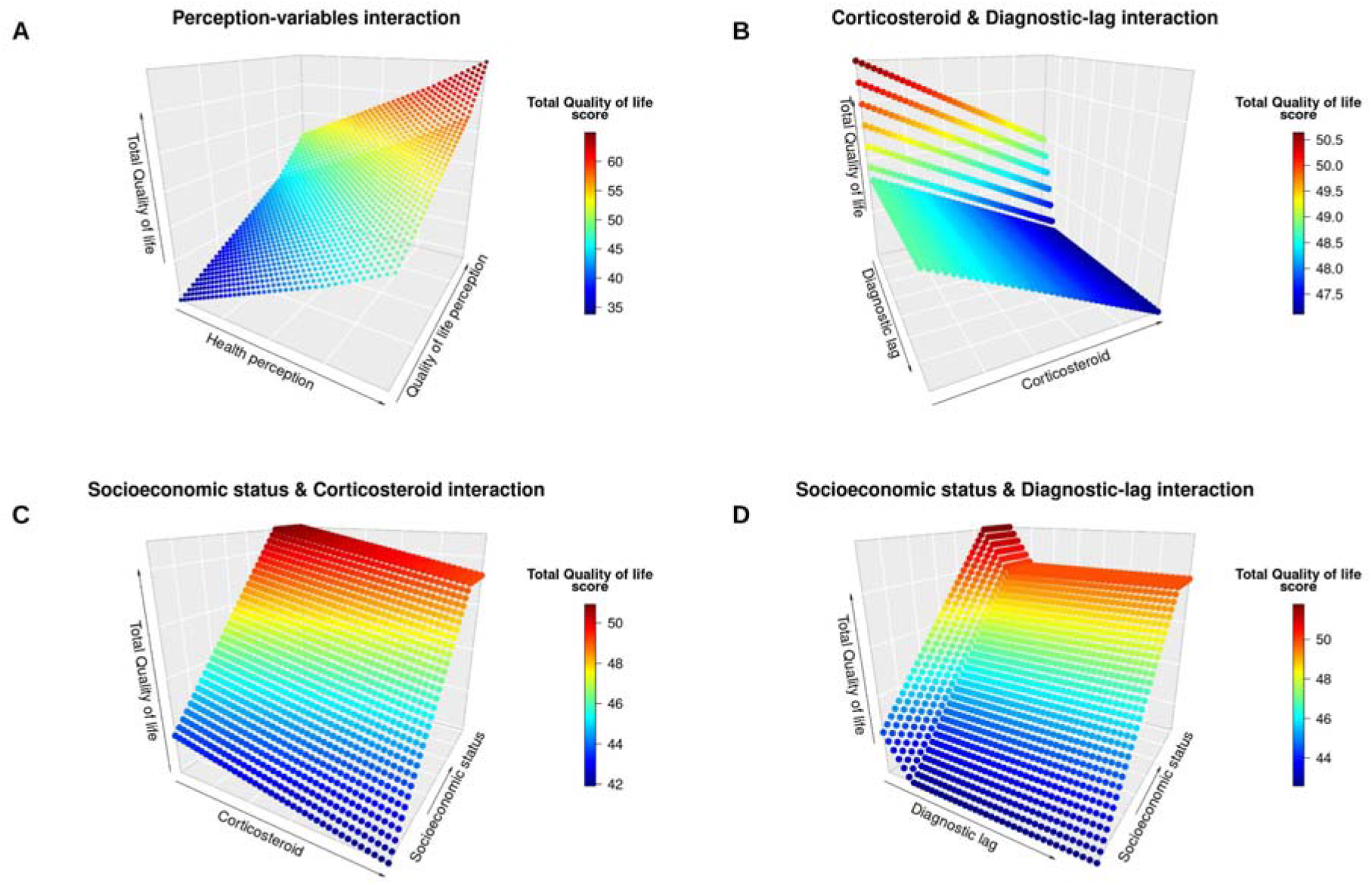
Dual-interaction partial dependence plots. Panel A showed the strongest effect over WHOQoL, which derived from health-perception and QoL-perception combination. Panel B and panel D show how early SLE diagnosis diminishes the negative effect associated with taking corticosteroids and with lower socioeconomic levels; whereas, from panel C we appreciate how socioeconomic status almost vanishes the effect of taking corticosteroids.

## 4. DISCUSSION

Medical registries and population-based cohorts have allowed us to deepen into the variability of SLE manifestations among different populations, highlighting the increased risk and worst outcome in ethnic minorities, such as Latin Americans. Although there have been some previous efforts to identify the characteristics of Latin American people with SLE, the heterogeneity of our populations makes it necessary to establish national approaches. Here we describe the development of the Mexican Lupus Registry (Lupus RGMX), a database collection platform that focuses exclusively on the characteristics of the Mexican individuals with SLE. This first cohort analysis provided an insight into clinical and sociodemographic characteristics of Mexican people with SLE and how these factors contribute to their quality of life.

Until January 2023, the Lupus RGMX platform collected the registries of 1172 Mexican individuals with SLE (**Supplementary Table S3**). Ninety four percent of the cohort consisted of women, a slightly higher percentage than the 88% observed in the GLADEL cohort (B. A. Pons-Estel et al., 2004). Mean age at diagnosis was 27 years; earlier than in other cohorts as the GLADEL and the RELESSER, which mean ages at diagnosis were 30 and 33 years old, respectively (Gómez-Puerta et al., 2021; B. A. Pons-Estel et al., 2004).

In our study, registrations from the 32 states of México were obtained (**Figure 2**). Since some states exhibited a lower number of registries, we decided to look up the rheumatologists enrolled at the Mexican College of Rheumatology and observed a similar geographic distribution of rheumatologists than the registered participants (**Supplementary Figure S1**). In México, it has been calculated that there is 0.55 certified rheumatologist per 100,000 inhabitants, with a surfeit of specialists in the capital and metropolitan areas of the country, which is in line with the observed in other Latin American countries (Pineda et al., 2019; Rocha, 2017). The low proportion of rheumatologists per patient, along with the uneven distribution of these specialists across México, can lead to unequal access to health and medical services (Martínez-Martínez & Rodríguez-Brito, 2020). Further analysis is needed to determine if the low number of registries can be attributable only to the low availability of specialists, and if some other contributing factors need to be analyzed. Additional efforts should be made to reach more individuals, such as social media campaigns and formal collaborations with lupus patients associations and certified rheumatologists; especially, to collaborate through private hospitals to reach patients from the highest socioeconomic levels, which were not represented in the present cohort.

Social factors, such as socioeconomic status, are determinants for the accessibility to healthcare services, and have been previously proposed as key factors contributing to SLE outcome in Latin-American subjects (Pons-Estel et al., 2012). To estimate socioeconomic status of the cohort we applied the NSE 2022 rule from the AMAI (NSE, 2022); which established that Lupus RGMX cohorts belongs to the lower five socioeconomic levels of the seven included, with an estimated maximum monthly household mean income of 19,900 MXN (approximately 990 USD), of which between 35 to 54% is destined only to food and the rest has to cover communication, services, transport, education and healthcare (**Figure 3**). In México, a lower socioeconomic level is related to lower possibilities to access to healthcare; which can be translated into challenges to access medical specialists, longer waiting times for evaluations and follow-ups, and therefore, difficulties to obtain an early and accurate diagnosis; besides, they might experience shorter duration of appointments (due to saturation) and low availability of prescribed drugs, which many times are too expensive and difficult to obtain (Pineda et al., 2019). Further analysis will be needed to deepen the health disparities among Mexican people with SLE and the consequences of this inequality in their prognosis.

In complex diseases such as SLE, the heterogeneity in manifestations may complicate an early and accurate diagnosis; this delay not only in diagnosis, but also in further treatment applications, can contribute to a continuous inflammatory state that may potentially lead to damage; patients with early diagnosis have shown lower incidence rate of flares than those with late diagnosis (Oglesby et al., 2014). In consonance with this, the timely identification of clinical manifestations and further prevention of damage development is key to ensure better outcomes; in this context, self-reported surveys focused on clinical manifestations have been proposed as a potential approach to improve the communication between patient-clinician, and therefore, improve and enhance the early identification of clinical signs (Munroe et al., 2022). To determine the frequency of clinical manifestations, the Lupus RGMX registry included a questionnaire that considers the most common clinical manifestations in SLE patients, which allowed to identify which are most prevalent among the cohort, along with which manifestations are most likely to appear simultaneously, such as arthritis and lupus headache (**Figure 4**), which is in line with previous reports of joint complaints affecting up to 90% of people with SLE (Ceccarelli et al., 2022). Since it is a self-reported questionnaire, it also provides an insight on how the individual with SLE perceives their own health, which can be a useful tool to identify the main affections and necessities of this population. Lupus RGMX future efforts will focus on establishing collaborations with clinicians to evaluate the participants, and to provide a clinically valid indicator of their manifestations, such as the SLEDAI or the SLICC index, to compare with the observations established by the self-reported survey. The validation of the relevance and potential applications of self-reported clinical surveys will provide a tool not only for the prevention of SLE associated complications, but also an easier and more reachable approach to follow up the response to treatments (Tiwari et al., 2022).

Due to its importance in patient evolution, it has been recently proposed that, the self-perceived QoL of the individual with SLE should be considered in the design and decision-making of treatments; we therefore integrate the WHOQoL-bref in our platform (Elera-Fitzcarrald et al., 2018; Kernder et al., 2020). The Lupus RGMX cohort exhibited a significantly lower total WHOQoL-bref score than the comparison groups, in all domains, which is in line with previous reports and has been associated with the disease burden, pain, anxiety, fatigue, and other clinical manifestations (**Figure 5 and Figure 6**). Deepening into social and clinical variables that may be influencing QoL in our sample, we observed that significant differences in WHOQoL total scores were exhibited between different socioeconomic levels (**Figure 7**); with the lowest mean score in the E socioeconomic level (Monthly household income 261 USD, MXN 4,833). This association between socioeconomic level and QoL may be related to different aspects of the daily life of the people with SLE: factors such as retaining employment after the diagnosis, the possibility of access to health insurance or afford treatments that are not available through social health services, as well as high cost of additional medical health care, can lead to an emotional stress for the person with SLE, triggering mental health manifestations such as fatigue, anxiety and depression, which have been associated with a diminished quality of life (Hohls et al., 2021; Phuti et al., 2018).

According to the WHO, QoL is the perception an individual has about his position in life, and, in this self-perception, it is important to consider all the subject’s life context; in SLE it has been reported that sociodemographic factors such as educational level or socioeconomic status, as well as the clinical manifestations, influence the QoL of the individual (Pereira et al., 2020). The multivariate adaptive regression splines model allowed us to deepen into the effects and contributions of different variables over the total WHOQoL score in our population. Health perception, QoL perception and socioeconomic status were the variables with higher relative importance for our model, followed by corticosteroids consumption and the diagnosis lag (**Figure 8**). As previously mentioned, earlier diagnosis of SLE has been associated with a timely initiation of treatment, which may lead to decrease in the risk of flares and, therefore, lower tissue and organ damage, better outcome, and lower mortality rates. This improvement on the prognosis is also associated with a lower use of healthcare resources and infrastructures, such as hospitalization, consultation with specialists on private practice and treatments, which translates in lower healthcare costs (Oglesby et al., 2014). Over 90% of our cohort reported the consumption of glucocorticoids, the standard of care in SLE along with antimalarials due to its anti-inflammatory effects. Although effective, the use of glucocorticoids has been related with neuropsychiatric manifestations, such as depression and anxiety, previous studies have observed an inverse association between glucocorticoid dose and emotional health. It is also important to highlight that some of the side effects of their use, besides potential organ damage, are weight gain and other appearance-related effects, which can contribute to self-perception and mental health (Miyawaki et al., 2021; Porta et al., 2020). The sum of all these factors, in addition to physical limiting manifestations such as arthritis or lupus headache, influence the self-perception that the patient has over her health and QoL; which explain why self-perception of health and quality of life are the main variables contributing to QoL in our cohort. To date, correlation between patient own health perception and clinician perception has not been established, still, recent works highlight the importance of considering the patient’s perception, as well as mental health related manifestations, as this can bring under detected needs to the conversation and also have a positive impact, not only in their QoL, but also in other aspects such as adherence to treatment and self-management of the disease (Dima et al., 2019; Miyawaki et al., 2021; Porta et al., 2020).

Here, we showed a preliminary and descriptive analysis of the Lupus RGMX data. Limitations of this study and analysis include the low participation of individuals from some states in México, alongside with the accessibility of the Registry platform, due both socioeconomic status and internet availability, and the self-reported nature of the data. Despite this cohort do not include individuals from the highest socioeconomic status, the A/B and C+ levels, our model allows us to infer that, in these individuals, socioeconomic status may not be influencing their quality of life more than level D+ influences (**Figure 4 and Figure 9C**); further analysis will need to deepen the impact of socioeconomic status over other aspects of daily life of people with SLE. As for the lack of access to the internet by certain segments of the population, the prevalence of individuals among the lowest socioeconomic levels gives us a preliminary idea of how this population is being affected by SLE. Still, these limitations represent an opportunity to improve the quality of the data and to enrich its analysis, current efforts are focused on establishing collaboration with rheumatologists across the country, who invite and help the people with SLE to register on their clinical practice, to ensure a more complete and representative participation of Mexican people with SLE. Our regression model also presents a limitation: when two variables exhibit similar values, it chooses only one of the variables to be presented, this could be masking some variables, but this usually happens with variables that are assessing the same factor but with different units so, the general result should not change.

The strengths of this study include the interdisciplinary and synergistic work between clinicians, researchers, and people with SLE communities which through the creation and implementation of the Lupus RGMX have established a pioneering approach that will provide a reliable source of data of the medical history, prevalence, and clinical characteristics of SLE among the Mexican population. The consideration and understanding of the self-experience of the individual with SLE, in addition to the knowledge and advances of clinicians and researchers, will allow us to design strategies that lead to the improvement of the diagnosis, prognosis, and treatment of Mexican individuals with SLE.

This work observed a lower QoL among Mexican individuals with SLE when compared with non-SLE affected individuals, and that this can be explained in part by low incomes, diagnosis lag, and glucocorticoids consumption; the convergence of these factors promotes a lower self-perception of health and QoL among our cohort. Furthermore, self-perception factors, i.e., perceived health and perceived quality of life, better predict total WHOQoL, which highlighted the importance of considering patients’ perception for diagnosis, prognosis, and treatment. The Lupus RGMX platform has proven to be a helpful tool to provide information about SLE in México, its strengthening will potentially help to enrich the knowledge we have about how SLE affects our population.

## Data Availability

All data produced in the present study are available upon reasonable request to the authors

## Acknowledgments

This work received support from Luis Aguilar, Alejandro León, and Jair García of the Laboratorio Nacional de Visualización Científica Avanzada. We also thank Carina Uribe Díaz, and Alejandra Castillo Carbajal for their technical support.

