## Supplemental figures for "Lupus RGMX: Social and Clinical Characteristics and their Contribution to Quality of Life in a Mexican Cohort with SLE"

**SUPPLEMENTARY MATERIALS**

**Table S1.** Sociodemographic characteristics of SLE participants (n=1214) and non-SLE groups (n=687).

| | SLE patients (n=1214) | Controls from Lupus RGMX (n=179) | Controls from Twins MX (n=508) |
|---|---|---|---|
| Sex (n, %) | | | |
| Women | 1132 (93.2%) | 132 (73.7%) | 395 (77.8%) |
| Men | 72 (6.8%) | 47 (26.3%) | 113 (22.2%) |
| Mean age (SD) | 36.3 (10.9) | 30.9 (11.1) | 30.7 (9.5) |
| Socioeconomic status (%) | | | |
| E (261 USD) | 6.6% | 4.3% | - |
| D (430 USD) | 42.4% | 33.6% | - |
| D+ (595 USD) | 38.9% | 45.7% | - |
| C- (251 USD) | 9.8% | 7.1% | - |
| C (962 USD) | 2.3% | 9.3% | - |

**Table S2.** Variables feeding the multivariate adaptive regression splines model. Total-WHOQoL is the target variable, whereas the rest of them are predictor variables. All categorical variables were transformed to dummy variables by one-hot encoding technique.

| **Variable** | **Description** | **Range / levels** |
|---|---|---|
| Total WHOQoL (* this is the target variable) | Aggregated score of WHOQoL. | {17.83 : 75.62} |
| Overall quality of life perception | Self reported quality of life perception. | {1, 2, 3, 4, 5} |
| General health perception | Self reported general health perception. | {1, 2, 3, 4, 5} |
| Socioeconomic status | Socioeconomic score | {5 : 143} |
| Calculated age | Automatically calculated based on birthday. | {11 : 79} |
| Sex | Self reported sex | {1 = female,<br>0 = male} |
| Years living with SLE | Indicates how long from the first official SLE diagnostic. | {0 : 40} |
| Prednisolona | If true, indicates what corticosteroid takes. | {0 = Prednisone,<br>1 = Prednisolone,<br>2 = Deflazacort,<br>3 = Meticorten,<br>4 = Methylprednisolone,<br>5 = Calcort,<br>6 = Betamethasone,<br>7 = No,<br>8 = Other} |
| Treatment | Indicates what medications a patient takes regularly to treat SLE. | {0 = Antimalarial,<br>1 = Corticosteroid,<br>2 = Mycophenolic acid,<br>3 = Azathioprine,<br>4 = Methotrexate,<br>5 = Rituximab,<br>6 = Ciclosporin,<br>7 = Cyclophosphamide,<br>8 = Other treatment} |
| Comorbidity | Indicates co-occurring clinical conditions in SLE patients. | {1 = Diabetes,<br>2 = Hypertension,<br>3 = Cancer, |

| | | |
|---|---|---|
| | | 4 = Cardiovascular disease,<br>5 = Osteoporosis,<br>6 = Rheumatoid arthritis,<br>7 = Multiple sclerosis,<br>8 = Thyroid disease,<br>9 = Other comorbidity,<br>10 =No comorbidity} |
| Health provider | Indicates current medical services provider. | {1= Instituto Mexicano del Seguro Social (IMSS),<br>2= Instituto de Seguridad y Servicios Sociales de los Trabajadores del Estado (ISSSTE),<br>3 = Petróleos Mexicanos (PEMEX),<br>4 = Secretaría de la Defensa (SEDENA),<br>5 = Secretaría de Marina (SEMAR),<br>6 = Secretaría de Salud (SSa),<br>7 = Servicios Estatales de Salud (SESA),<br>8 = IMSS-Bienestar (sin seguridad social),<br>9 = Private} |
| Diagnostic lag | Indicates the time-lapse between first SLE symptom and the official clinical diagnostic. | {0, 1, 2, 3, 4, 5, 6, 7} |

**Table S3**. Socio-demographic characteristics of the participants with SLE in Lupus RGMX (n=1172).

| | SLE patients (n=1172) |
|---|---|
| Sex (n, %) | |
| Women | 1101 (93.9%) |
| Men | 71 (6.1%) |
| Age (SD) | 36.6 (10.7) |
| Occupation | |
| Student | 135 (11.5%) |
| Employee | 485 (41.4%) |
| Retired | 47 (4.0%) |
| Unemployed | 312 (26.6%) |
| Unspecified | 193 (16.5%) |
| Education | |
| None | 2 (0.2%) |
| Elementary school (9 years) | 5 (0.4%) |
| Junior high school (12 years) | 76 (6.5%) |
| Senior high school/Technician (15 years) | 310 (26.5%) |
| College | 442 (37.7%) |
| Graduate school | 144 (12.3%) |
| Unspecified | 193 (16.5%) |
| Time since diagnosis (SD) | 8.6 (7.9) |
| Pregnant participants (n,%) | 9 (0.8%) |
| Familiar with SLE (n,%) | 211 (18.0%) |
| Nephritis (n, %) | 263 (22.4%) |
| Health service provider (n, %) | |
| IMSS | 551 (47.0%) |
| Private | 351 (29.9%) |
| ISSSTE | 114 (9.7%) |
| SSa | 87 (7.4%) |
| IMSS-bienestar | 27 (2.3%) |
| PEMEX | 13 (1.1%) |
| Other | 9 (0.8%) |
| SESA | 9 (0.8%) |
| SEDENA | 6 (0.5%) |
| SEMAR | 5 (0.4%) |
| Treatment (n, %) | |
| Glucocorticoids | 838 (91.93%) |

| | |
|---|---|
| Antimalarials | 192 (21.1%) |
| Mycophenolate | 85 (9.3%) |
| Azathioprine | 67 (7.3%) |
| Methotrexate | 47 (5.2%) |
| Rituximab | 14 (1.5%) |
| Cyclosporine | 4 (0.4%) |
| Cyclophosphamide | 5 (0.5%) |
| Others | 67 (3.4%) |

IMSS: Instituto Mexicano del Seguro Social; ISSSTE: Instituto de Seguridad y Servicios Sociales de los Trabajadores; SSa: Secretaría de Salud; PEMEX: Petróleos Mexicanos; SEDENA: Secretaría de la Defensa Nacional; SESA: Servicios Estatales de Salud; SEMAR: Secretaría de la Marina.

**Table S4**. Number of registered participants and Board-Certified rheumatologists (according to the Mexican College of Rheumatology) in 32 states of Mexico.

| State | Registered participants | Rheumatologists in the State | Total population, 2020 |
|---|---|---|---|
| Aguascalientes | 3 | 8 | 1,425,607 |
| Baja California | 17 | 22 | 3,769,020 |
| Baja California Sur | 2 | 2 | 798,447 |
| Campeche | 9 | 2 | 928,363 |
| Coahuila | 9 | 16 | 3,146,771 |
| Colima | 2 | 3 | 731,391 |
| Chiapas | 14 | 9 | 5,543,828 |
| Chihuahua | 23 | 14 | 3,741.869 |
| México City | 235 | 152 | 9,209,944 |
| Durango | 10 | 4 | 1,832,650 |
| Guanajuato | 35 | 15 | 6,166,934 |
| Guerrero | 12 | 3 | 3,540,685 |
| Hidalgo | 27 | 4 | 3,082,841 |
| Jalisco | 27 | 42 | 8,348,151 |
| State of México | 131 | 42 | 16,992,418 |
| Michoacán | 22 | 11 | 4,748,846 |
| Morelos | 60 | 6 | 1,971,520 |

| | | | |
|---|---|---|---|
| Nayarit | 6 | 1 | 1,235,456 |
| Nuevo León | 29 | 38 | 5,784,442 |
| Oaxaca | 12 | 9 | 4,132,148 |
| Puebla | 46 | 17 | 6,583,278 |
| Querétaro | 107 | 20 | 2,368,467 |
| Quintana Roo | 14 | 3 | 1,857,985 |
| San Luis Potosí | 16 | 13 | 2,822,255 |
| Sinaloa | 8 | 11 | 3,026,943 |
| Sonora | 15 | 12 | 2,944,840 |
| Tabasco | 9 | 5 | 2,402,598 |
| Tamaulipas | 10 | 6 | 3,527,735 |
| Tlaxcala | 15 | 1 | 1,342,977 |
| Veracruz | 36 | 10 | 8,062,579 |
| Yucatán | 33 | 16 | 2,320,898 |
| Zacatecas | 9 | 4 | 1,622,138 |
| TOTAL | 1172 | 521 | |

**Table S5.** Results of WHOQoL comparison by permutation test for all QoL dimensions.

| Comparison | Expected difference | p-value | SE |
|---|---|---|---|
| Environmental, RedCap - SLE = 0 | 1.12 | 0.011 | 0.33 |
| Environmental, Twins - SLE = 0 | 2.10 | 0.000 | 0.32 |
| Physical, RedCap - SLE = 0 | 2.89 | 0.000 | 0.32 |
| Physical, Twins - SLE = 0 | 4.30 | 0.000 | 0.33 |
| Psychological, RedCap - SLE = 0 | 1.61 | 0.002 | 0.39 |
| Psychological, Twins - SLE = 0 | 2.18 | 0.000 | 0.41 |
| Social, RedCap - SLE = 0 | 2.13 | 0.001 | 0.47 |
| Social, Twins - SLE = 0 | 2.01 | 0.004 | 0.48 |
| Total QoL, RedCap - SLE = 0 | 7.67 | 0.000 | 1.26 |
| Total QoL, Twins - SLE = 0 | 10.49 | 0.000 | 1.21 |

**Table S6.** Results of WHOQoL comparison by pairwise permutation test across all socioeconomic levels. P-values were corrected by false discovery rate.

| Comparison | Expected difference | p-value | p-adjust |
|---|---|---|---|
| C - C- = 0 | 1.49 | 0.137 | 0.153 |
| C - D = 0 | 4.08 | 0.000 | 0.000 |
| C - D+ = 0 | 2.02 | 0.044 | 0.062 |
| C - E = 0 | 4.36 | 0.000 | 0.000 |
| C- - D = 0 | 4.48 | 0.000 | 0.000 |
| C- - D+ = 0 | 0.73 | 0.464 | 0.464 |
| C- - E = 0 | 4.29 | 0.000 | 0.000 |
| D - D+ = 0 | -5.88 | 0.000 | 0.000 |
| D - E = 0 | 1.94 | 0.053 | 0.066 |
| D+ - E = 0 | 4.82 | 0.000 | 0.000 |

**Figure S1**. Number of registered participants and Board-Certified rheumatologists (according to the Mexican College of Rheumatology) in 32 states of Mexico.

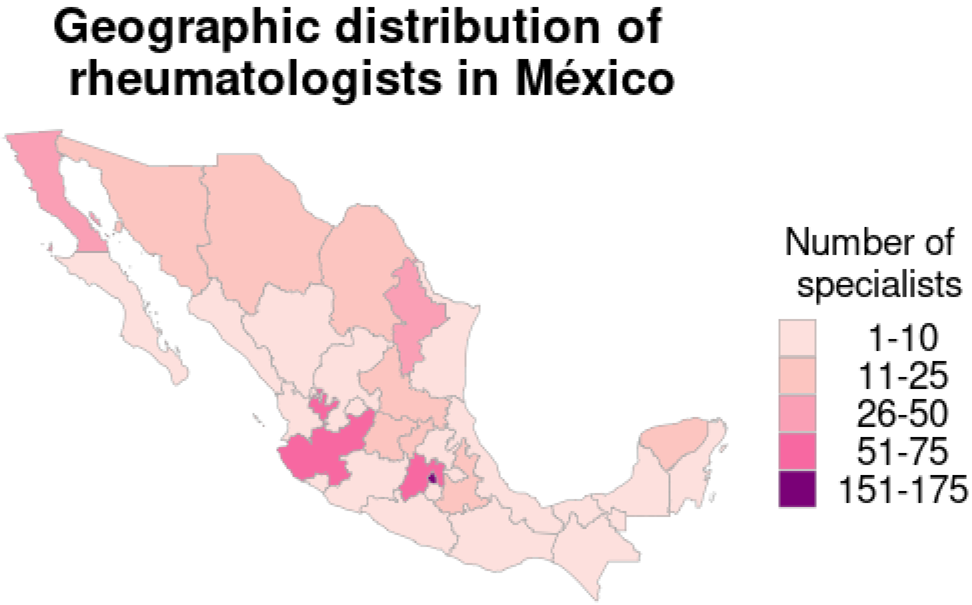
